# Time needed to perform intermittent catheterization in adults with spinal cord injury: A pilot randomized controlled cross-over study

**DOI:** 10.1101/2021.08.16.21253936

**Authors:** Karthik Gopalakrishnan, Nick Fabrin Nielsen, Andrea L. Ramirez, Jeppe Sørensen, Matthias Walter, Andrei V. Krassioukov

**Author notes:** **Corresponding author:** Andrei V. Krassioukov MD, PhD, FRCPC, International Collaboration on Repair Discoveries, Blusson Spinal Cord Centre, 818 West 10th Avenue, Vancouver, British Columbia V5Z 1M9, Canada. These authors share the senior authorship (Matthias Walter, Andrei V. Krassioukov).

## Abstract

**Background:** Intermittent catheterization (IC), considered the gold standard for bladder management for individuals with spinal cord injury (SCI) with sufficient dexterity, is usually performed using hydrophilic (HPC) or non-hydrophilic (non-HPC) catheters. Currently, there is no evidence on the temporal burden associated with IC with either catheter.

**Objective:** To compare both catheters regarding their time requirement for IC and participant satisfaction.

**Design, setting and participants:** Twenty individuals with chronic (>1-year) SCI at any spinal segment were randomized to undergo two cross-over assessments within 10 days (i.e., either starting with HPC or non-HPC). We measured time taken to perform IC using a 13 step pre-determined IC protocol (e.g., enter bathroom, wash hands, transfer to toilet, etc.). Furthermore, we assessed user satisfaction of both catheters using a Likert scale (i.e., strongly agree=5, strongly disagree=1).

**Outcome measures and statistical analysis:** Time (i.e., for each step and in total) to perform IC and participant satisfaction were compared between catheters using non-parametric statistics, i.e., Wilcoxon rank sign tests. Results are presented as median with interquartile range.

**Results and limitations:** Participants using HPCs spent less time to prepare a catheter [15 s (10-20) vs. 41 (20-69), p=0.002] and overall to perform IC [283 s (242-352) vs. 373 (249-441), p=0.01] compared to non-HPCs. Moreover, participants rated the preparation of HPCs to be easier [5 (4-5) vs. 4 (2-4), p=0.047] compared to non-HPCs. The key limitation of this pilot study was the sample size.

**Conclusions:** Preparation and usage of HPCs for IC is easier and faster compared to non-HPCs. IC can be a significant temporal burden for SCI individuals.

**Patient summary:** We compared coated and uncoated catheters on time needed for intermittent catheterization and user satisfaction in individuals with spinal cord injury. Participants can manually empty their bladder quicker and easier with coated compared to uncoated catheters.

## INTRODUCTION

Spinal cord injury (SCI) poses a significant burden on affected individuals and their quality of life (QoL).^1^ Adult neurogenic lower urinary tract dysfunction (ANLUTD) is a common complication resulting from damage to the sensorimotor and autonomic nervous system^2^, entailing urinary storage and/or voiding dysfunctions potentially jeopardizing the entire urinary tract if left untreated.^3^

Intermittent catheterization (IC) is considered the gold standard for bladder management for individuals with SCI with sufficient dexterity^4^. There are two main types of catheters for IC. Hydrophilic catheters (HPC) have a polymer coating which binds to water^5^ leading to a smooth and slippery surface. HPCs do not require manual lubrication and are usually pre-packaged in sterile water.^6^ In contrast, uncoated non-hydrophilic catheters (non-HPC) require manual lubrication to reduce insertion-related friction.^7^ Over the last decade, HPCs have been associated with decreased incidence of urinary tract infections (UTIs) compared to non-HPCs^8^, but considering the heterogeneity with respect to the definition of UTIs in these studies^9^, definitive comparisons between catheter types regarding the incidence of related UTIs are still pending.^10^

The slippery surface of HPCs appears to reduce friction as well as the incidence of urethral injuries.^11^ Furthermore, pre-lubrication makes HPCs easier to use because manual lubrication is not needed, which supports individuals with impaired dexterity.^12^

Given these benefits, one might assume that HPCs would be the prevailing catheter type. In 2008, Woodbury et al. reported that 74% of Canadian IC-users utilized non-HPCs, while only a minority used HPCs (15%) or a combination of both (11%).^13^ Since HPCs are more expensive than non-HPCs (both inside and outside of North America)^14,15^ and most Canadian provinces provide only limited support for intermittent catheters^16^, SCI-induced financial hardship makes it difficult to pay for HPCs out-of-pocket.^17^

With a recommended IC frequency of 4 to 6 times per day^18^ and the consequent total time requirement, time needed to perform IC remains a paramount aspect of the catheterization process. The significant IC time requirement can be taxing on personal relationships and may reduce available leisure time thereby resulting in inter- and intrapersonal strains. Further, it may be a disadvantage in the workforce due to out-of-home IC challenges, such as small toilets and hygiene issues.^19^ In addition, there may be a burden associated with self-catheterization that could be exacerbated by a perceived need to carry equipment such as wipes and hand sanitizer in order to perform IC outside the home setting. Other challenges of IC include opening the catheter packaging, preparing and inserting the catheter for catheterization, particularly for those with impaired dexterity.^20,21^

Considering the lack of evidence on whether HPCs or non-HPCs are more efficient with respect to time required for bladder management and the aforementioned characteristics and limitations, our aim was to compare both types of catheters regarding time needed to perform IC and user satisfaction.

## MATERIALS AND METHODS

### Ethics, study design and participants

This pilot cross-over randomized controlled trial (RCT) was approved by the University of British Columbia (UBC) Research Ethics Board (H17-03228) and Vancouver Coastal Health (V17-03228), and registered at clinicaltrials.gov (NCT05003999), while conforming to the Declaration of Helsinki. Our target number of participants (n = 20) for this pilot cross-over RCT is in line with general recommendations from the literature.^22,23,24^ Sixty-five individuals were screened between December 2018 and April 2019 according to the inclusion and exclusion criteria from which 21 individuals were invited for a baseline visit (Fig. 1, the consort flow diagram)^25^. The criteria were as follows: inclusion – female or male, age 18 years or older, presenting with chronic (>1-year post-injury) SCI at any spinal level, hand function sufficient to perform IC, fluent in English, not being pregnant, and without history of urinary diversion; exclusion – acute medical issues that would adversely affect participation in the study and members of the investigational team or their immediate family. After obtaining informed consent, 20 individuals were assigned a unique number during the baseline visit. Furthermore, neurological level and completeness of injury were classified in accordance with the International Standards for Neurological Classification of SCI (ISNCSCI)^26^ by a physician generating a grade on the American Spinal Injury Association impairment scale (AIS). Urine samples were collected to exclude or confirm UTIs (i.e., urine cultures and clinical symptoms)^27^ and in all female participants to confirm the absence of pregnancy. One eligible individual withdrew due to personal reasons, who was replaced by an eligible backup (i.e. screening individual #21). After enrollment into the study, participants were randomized into two groups (i.e., starting with an HPC or non-HPC, allocation ratio 1:1) using a random sequence generator. Thereafter, participants underwent the first assessment followed by the second cross-over assessment within 10 days. Participants were followed up 5 to 10 days later via phone regarding potential adverse events (AEs). All catheters were provided by Coloplast A/S (Humlebæk, Denmark), i.e., HPC (SpeediCath®) and non-HPC (Self-Cath®). For non-HPC, lubrication jelly (MUKO®, 3.5g package, Cardinal Health Canada Inc, Toronto, Canada) was provided.

**Figure 1–.**
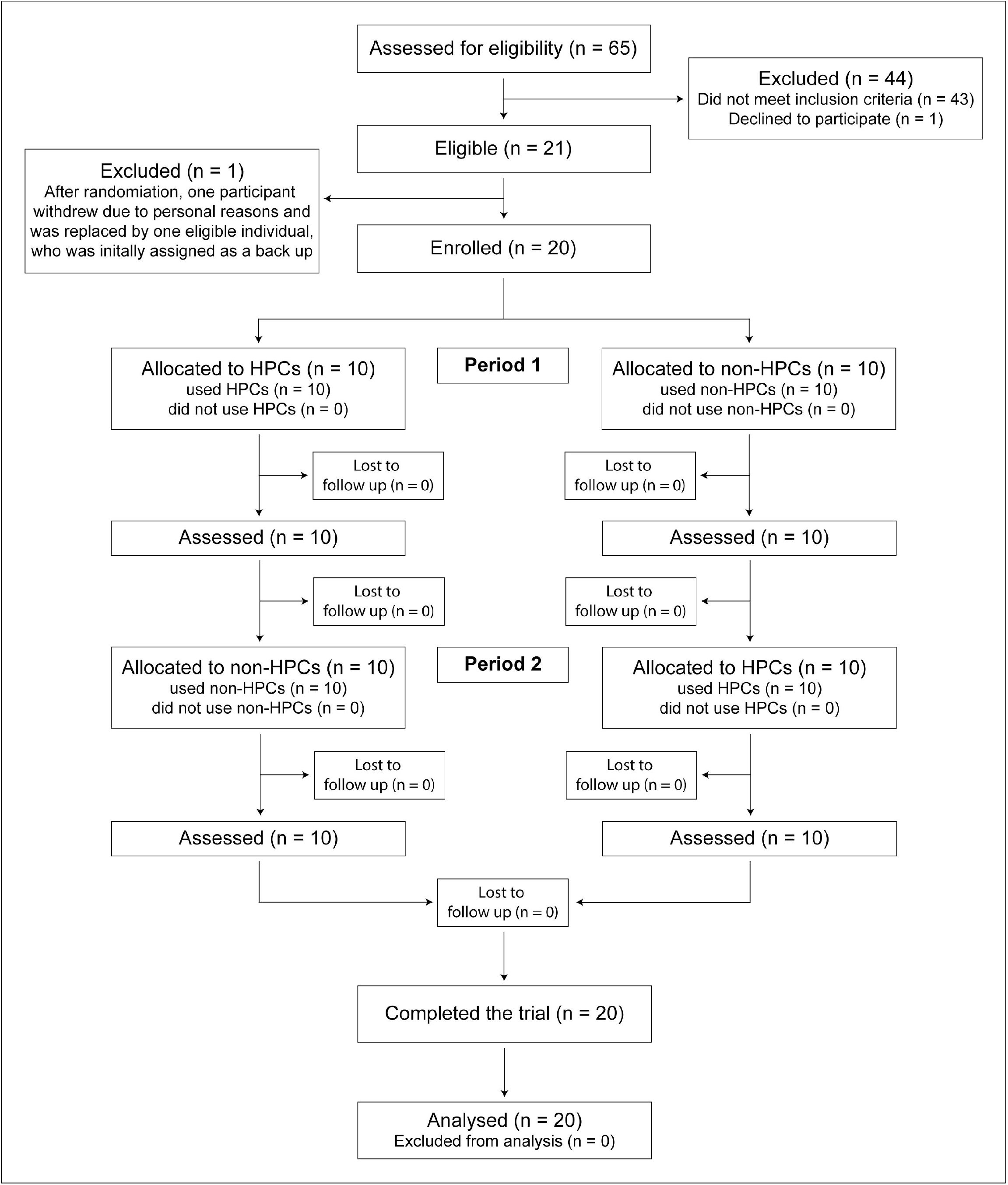
The Consort 2010 Flow Diagram. HPC = hydrophilic catheter, non-HPC = non-hydrophilic catheter

### Primary and secondary outcome objectives

The two primary outcome objectives of this study were the time needed to perform IC and user satisfaction (between HPC and non-HPC). The secondary outcome objectives were time needed to perform IC with respect to the participants’ level of injury and catheter use in daily life.

### Assessment visits

All assessments were performed at UBC, Vancouver, Canada. A predetermined 13 step IC protocol was drafted together with individuals with SCI performing IC (Table 2). At each assessment visit, participants performed a guided test run to familiarize themselves with the protocol. Then, each step was recorded separately. To ensure discretion, one investigator was situated inside the bathroom behind a curtain. This investigator instructed the participant to “start” each step at which point the investigator outside started the timer. After completion of each step, the participant said “I’m done” and the timer was stopped. Subsequently, the participant moved on to perform the next step of the protocol. At the end of each assessment, participants completed a questionnaire (see supplementary Fig. 1 and 2) regarding their catheter satisfaction. At the end of the second visit, participants were also asked about their preference between the two catheters.

### Statistical analysis

Statistical analyses were conducted using STATA 15 (StataCorp, College Station, TX, USA). Data are presented as raw values and percentages. Results are presented as median with interquartile range (IQR). In addition, range (i.e., min - max) is provided for age and time post injury. Non-parametric statistics were conducted, i.e., the Wilcoxon signed-rank test for primary and secondary outcomes (i.e., all variables except for the preferred catheter question which was tested with the Wilcoxon rank-sum test). All statistical tests were two-sided, and p < 0.05 was considered statistically significant.

## RESULTS

### Participant characteristics

Twenty participants (five females) with a median age of 45 years (36 - 53, range 26 - 65) and median time post injury of 14 years (10 - 26, range 3 - 46) completed the study (Table 1). The majority were daily non-HPCs (75%, 15/20) users. One participant had a symptomatic UTI six days after the second assessment and was treated with antibiotics appropriately.

**Table 1–.** Participant demographics and injury characteristics.

| No. | NLI | AIS | Sex | Age range* [yrs] | TPI range* [yrs] | Catheter use in daily life | Catheter size<sup>§</sup> [Fr] |
| --- | --- | --- | --- | --- | --- | --- | --- |
| 1 | C5 | B | 1 | 36-40 | 6-10 | Non-HPC | 14 |
| 2 | C5 | B | 2 | 26-30 | 11-15 | HPC | 14 |
| 3 | C5 | B | 1 | 61-65 | 11-15 | HPC | 14 |
| 4 | C5 | B | 1 | 36-40 | 16-20 | HPC | 14 |
| 5 | C6 | A | 1 | 31-35 | 11-15 | Non-HPC | 16 |
| 6 | C6 | A | 2 | 41-45 | 21-25 | Non-HPC | 14 |
| 7 | C6 | B | 1 | 31-35 | 1-5 | Non-HPC | 14 |
| 8 | C6 | B | 1 | 31-35 | 6-10 | HPC | 14 |
| 9 | C6 | B | 1 | 51-55 | 21-25 | Non-HPC | 14 |
| 10 | T2 | A | 1 | 26-30 | 11-15 | Non-HPC | 14 |
| 11 | T2 | A | 1 | 51-55 | 36-40 | Non-HPC | 14 |
| 12 | T2 | B | 1 | 51-55 | 6-10 | Non-HPC | 14 |
| 13 | T3 | A | 1 | 41-45 | 26-30 | Non-HPC | 16 |
| 14 | T4 | A | 1 | 36-40 | 16-20 | Non-HPC | 14 |
| 15 | T4 | A | 2 | 41-45 | 26-30 | Non-HPC | 12 |
| 16 | T4 | A | 2 | 51-55 | 46-50 | Non-HPC | 14 |
| 17 | T5 | A | 1 | 46-50 | 1-5 | HPC | 14 |
| 18 | T12 | A | 2 | 51-55 | 31-35 | Non-HPC | 12 |
| 19 | T12 | B | 1 | 61-65 | 11-15 | Non-HPC | 12 |
| 20 | L2 | D | 1 | 51-55 | 11-15 | Non-HPC | 14 |

\* For information, such as age and time post injury, that would allow the study participant or their family, friends or neighbors to identify them, we chose to provide a 5-year range for age and time post injury, rather than revealing the actual numbers.
§ Size of catheter provided in the study was the same size as the participant's regular catheter.
AIS = American Spinal Injury Association Impairment Scale, C = cervical, Fr = French, HPC = hydrophilic catheter, L = lumbar, NLI = neurological level of injury, No. = number, non-HPC = non-hydrophilic catheter, TPI = time post-injury, T = thoracic; yrs = years.

### Time comparison between HPC and non-HPC

Table 2 highlights the time comparison between HPC and non-HPC for each of the 13 steps and in total. Total time taken to execute all 13 steps (i.e., from entering to exiting the bathroom) was 90 seconds shorter when using HPC compared to non-HPC [283 s (242 - 352) vs. 373 (249 - 441), p=0.01]. Total time before the actual use of a catheter (i.e., steps 1 to 5, table 2) was not statistically different between HPCs and non-HPCs [86 s (71 - 119) vs. 95 (74 - 121), p=0.296]. Similarly, there was no significant difference in total time taken post catheter use (i.e., steps 10 to 13, table 2) between HPCs and non-HPCs [88 s (69 - 110) vs. 88 (71 - 104), p=0.218]. However, the grouped steps involving catheter use (i.e., steps 6 to 9, table 2), showed that the HPC group spent significantly less time than the non-HPC group [119 s (73 - 133) vs. 160 (85 - 222), p=0.014]. Of the individual steps, only the time taken to prepare the catheter was statistically significantly shorter for HPC compared to non-HPC (p=0.002).

**Table 2–.** Time comparison between HPC and non-HPC.

| No. | Steps | HPC | non-HPC | p value |
| --- | --- | --- | --- | --- |
| 1 | Enter bathroom | 15 (12-17) | 16 (13-17) | 0.304 |
| 2 | Wash hands | 28 (23-35) | 34 (22-38) | 0.097 |
| 3 | Transfer to toilet | 9 (8-15) | 10 (7-12) | 0.455 |
| 4 | Undress | 14 (9-23) | 13 (9-27) | 0.823 |
| 5 | Clean genitalia | 29 (20-43) | 30 (22-48) | 0.657 |
| **Total time before catheter use** |  | **86 (71-119)** | **95 (74-121)** | **0.296** |
| 6 | Prepare catheter | 15 (10-20) | 41 (20-69) | **0.002** |
| 7 | Insertion | 28 (15-41) | 44 (15-66) | 0.279 |
| 8 | Emptying | 36 (17-65) | 43 (25-53) | 0.737 |
| 9 | Removal and disposal | 18 (11-23) | 22 (12-30) | 0.113 |
| **Total time during catheter use** |  | **119 (73-133)** | **160 (85-222)** | **0.014** |
| 10 | Dress up | 32 (14-41) | 31 (15-42) | 0.305 |
| 11 | Transfer to wheelchair | 5 (4-7) | 9 (8-11) | 0.18 |
| 12 | Wash hands | 33 (26-43) | 31 (25-43) | 0.502 |
| 13 | Exit bathroom | 20 (17-24) | 20 (16-23) | 0.794 |
| **Total time post catheter use** |  | **88 (69-110)** | **88 (71-104)** | **0.218** |
| **Total time needed to perform IC** |  | **283 (242-352)** | **373 (249-441)** | **0.010** |

HPC = hydrophilic catheter, IC = Intermittent catheterization, No. = number of steps, non-HPC = non-hydrophilic catheter.

### Participants’ satisfaction of HPC and non-HPC use

HPC use was associated with significantly higher scores for “easiness to prepare” [5 (4 - 5) vs. 4 (2 - 4), p=0.047] and “did not feel burning” [5 (4 - 5) vs. 4 (3 - 5), p=0.042] compared to non-HPC. There were no statistical differences in any of the remaining scores (Table 3).

**Table 3–.**
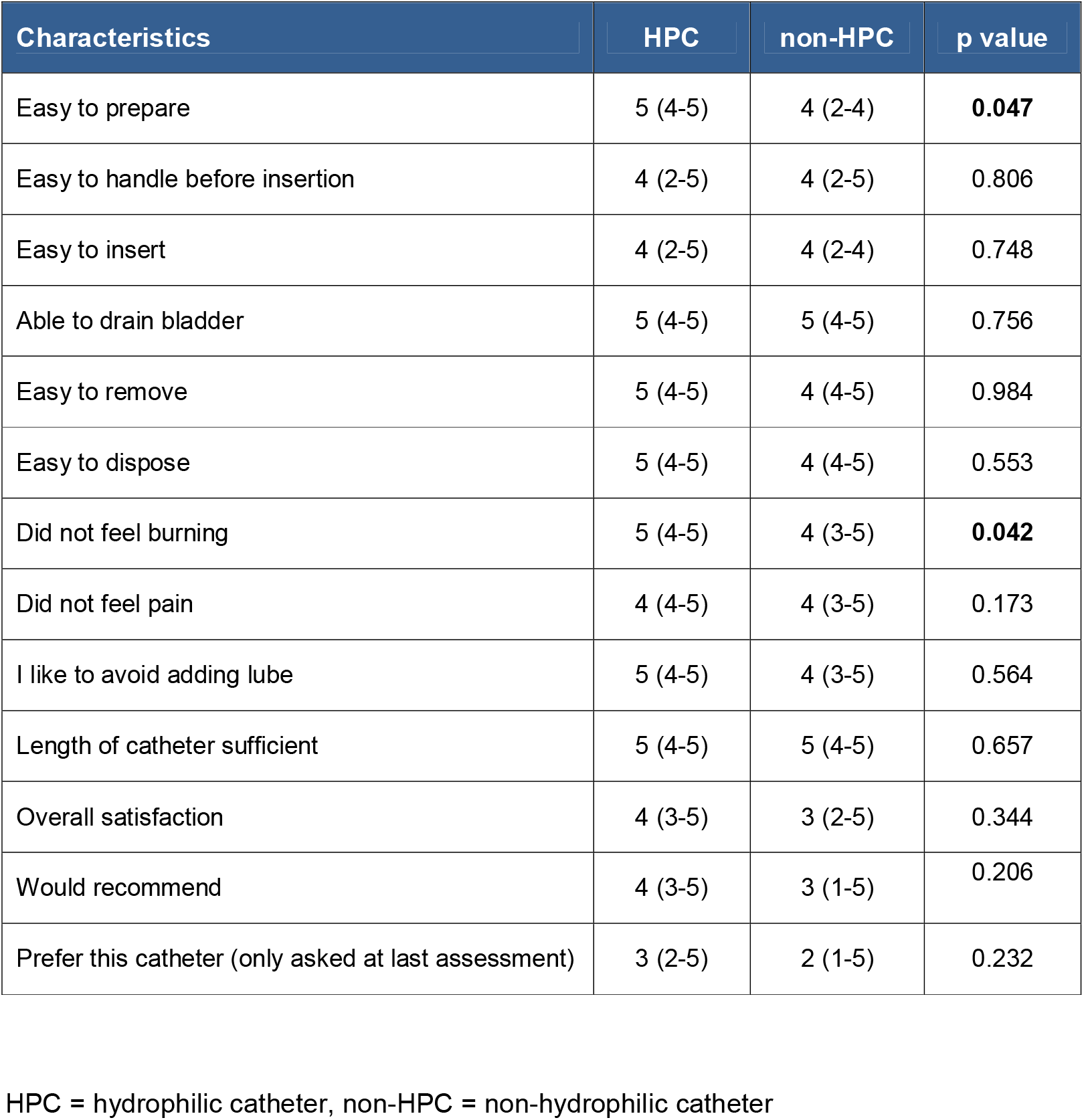
Comparison of participants’ satisfaction between HPC and non-HPC.

### Time comparison with respect to the participants’ level of injury and catheter use in daily life

For tetraplegic participants (9/20), total time taken to execute all 13 steps [300 s (249 - 352) vs. 421 (398 - 515), p=0.139], total time before using a catheter [89 s (78 - 107) vs. 99 (83 - 124), p=0.214], and total time of catheter use [119 s (71 - 160) vs. 221 (163 - 241), p=0.214] were all shorter when using a HPC compared to a non-HPC but did not yield statistical significance (table 4). Total time post catheterization was similar between HPCs and non-HPCs [94 s (71 - 108) vs. 92 (84 - 97), p=0.214]. For paraplegic participants (11/20), total time overall was significantly shorter when using HPC compared to a non-HPC [272 s (210 - 353) vs. 296 (225 - 433), p=0.041]. There was no significant difference between HPCs and non-HPCs for total time before [86 s (66 - 130) vs. 86 (65 - 119), p=0.657], during [118 s (80 - 124) vs. 128 (83 - 167), p=0.062] or post catheter use [78 s (69 - 105) vs. 71 (69 - 111), p=0.594].

**Table 4–.** Time comparison between HPC and non-HPC with respect to the participants’ neurological level of injury and type of catheter used in daily life.

| No. | Steps | Tetraplegia (n = 9) |  |  | Paraplegia (n = 11) |  |  | HPC-user (n = 5) |  |  | non-HPC user (n = 15) |  |  |
| --- | --- | --- | --- | --- | --- | --- | --- | --- | --- | --- | --- | --- | --- |
|  |  | HPC | non-HPC | p | HPC | non-HPC | p | HPC | non-HPC | p | HPC | non-HPC | p |
| 1 | Enter bathroom | 16 (16-20) | 16 (16-19) | 0.314 | 12 (11-15) | 13 (12-16) | 0.504 | 16 (16-21) | 18 (16-18) | 0.5 | 14 (11-16) | 15 (12-16) | 0.094 |
| 2 | Wash hands | 27 (23-33) | 34 (31-37) | 0.066 | 29 (24-35) | 29 (21-39) | 0.79 | 26 (22-27) | 34 (31-35) | 0.08 | 30 (24-36) | 34 (21-40) | 0.443 |
| 3 | Transfer to toilet † | 10 (8-25) | 11 (9-13) | 0.767 | 9 (8-10) | 9 (6-12) | 0.286 | 8 (7-9) | 9 (6-11) | 0.5 | 9 (8-25) | 11 (7-15) | 0.28 |
| 4 | Undress | 18 (12-41) | 17 (12-31) | 0.678 | 11 (9-18) | 10 (9-25) | 0.477 | 18 (12-34) | 16 (12-25) | 0.345 | 12 (9-20) | 11 (9-27) | 0.733 |
| 5 | Clean genitalia | 30 (25-34) | 28 (24-39) | 1.0 | 26 (19-47) | 34 (19-48) | 0.866 | 34 (25-42) | 32 (28-41) | 1.0 | 26 (19-43) | 28 (19-48) | 0.612 |
| **Total time before catheter** |  | 89 (78-107) | 99 (83-124) | 0.214 | 86 (66-130) | 86 (65-119) | 0.657 | 89 (75-131) | 119 (83-124) | 0.345 | 86 (68-107) | 92 (71-104) | 0.364 |
| 6 | Prepare catheter | 16 (11-20) | 67 (46-81) | **0.038** | 15 (9-24) | 21 (14-45) | **0.023** | 15 (11-17) | 46 (38-67) | **0.043** | 16 (10-24) | 36 (16-78) | **0.021** |
| 7 | Insertion | 32 (19-79) | 63 (55-124) | 0.441 | 26 (14-33) | 31 (13-51) | 0.534 | 19 (11-26) | 51 (22-63) | 0.225 | 30 (17-48) | 38 (13-70) | 0.57 |
| 8 | Emptying | 25 (16-32) | 27 (17-48) | 0.767 | 42 (30-67) | 45 (37-57) | 0.477 | 27 (25-32) | 22 (17-48) | 0.686 | 40 (16-67) | 45 (36-56) | 1.0 |
| 9 | Removal and disposal | 19 (15-32) | 25 (18-43) | 0.286 | 15 (8-22) | 16 (10-30) | 0.248 | 15 (13-19) | 25 (18-30) | 0.08 | 18 (8-32) | 18 (11-30) | 0.379 |
| **Total time during catheter** |  | 119 (71-160) | 221 (163-241) | 0.139 | 118 (80-124) | 128 (83-167) | 0.062 | 75 (61-119) | 201 (102-221) | 0.08 | 120 (80-158) | 158 (84-223) | 0.061 |
| 10 | Dress up | 32 (21-36) | 39 (15-42) | 0.678 | 32 (12-43) | 30 (15-42) | 0.197 | 36 (23-36) | 41 (39-43) | 0.225 | 32 (12-43) | 28 (11-42) | 0.755 |
| 11 | Transfer to wheelchair */** | N/A* | N/A* | N/A* | 5 (4-7) | 9 (8-11) | 0.18 | N/A** | N/A** | N/A** | 5 (4-7) | 9 (8-11) | 0.18 |
| 12 | Wash hands | 32 (26-39) | 32 (28-42) | 0.086 | 33 (23-49) | 27 (24-44) | 0.929 | 26 (25-32) | 28 (25-32) | 0.686 | 35 (30-49) | 40 (25-47) | 0.57 |
| 13 | Exit bathroom | 24 (18-26) | 21 (20-25) | 0.766 | 20 (14-20) | 18 (15-21) | 0.859 | 18 (17-26) | 21 (18-25) | 0.686 | 20 (14-24) | 20 (15-23) | 0.887 |
| **Total time after catheter** |  | 94 (71-108) | 92 (84-97) | 0.214 | 78 (69-105) | 71 (69-111) | 0.594 | 85 (71-108) | 97 (87-111) | 0.225 | 90 (69-105) | 84 (70-97) | 0.496 |
| **Total time needed to perform IC** |  | 300 (249-352) | 421 (398-515) | 0.139 | 272 (210-353) | 296 (225-443) | **0.041** | 249 (235-300) | 421 (241-491) | 0.08 | 285 (253-353) | 348 (257-433) | **0.047** |

†/\* Participants with tetraplegia did only transfer from the sink to the toilet (i.e. step 3) by wheeling but did not transfer from the wheelchair onto the toilet. They performed catheterization while sitting in their wheelchair. \*\* HPC-user were all participants with tetraplegia. HPC = hydrophilic catheter, IC = Intermittent catheterization, n = number of participants, N/A = not applicable, No. = number of steps, non-HPC = non-hydrophilic

For daily HPC users (5/20), total time overall [249 s (235 - 300) vs. 421 (241 - 491), p=0.08], before [89 s (75 - 131) vs. 119 (83 - 124), p=0.345], during [75 s (61 - 119) vs. 201 (102 - 221), p=0.08] and post catheter use [85 s (71 - 108) vs. 97 (87 - 111), p=0.225] were all shorter with HPC compared to non-HPC, but without significant differences (Table 4).

For daily non-HPC users (15/20), total time overall was significantly shorter with HPC compared to non-HPC [285 s (253 - 353) vs. 348 (257 - 433), p=0.047]. Total time before [86 s (68 - 107) vs. 92 (71 - 104), p=0.364] and during catheter use [119 s (71 - 160) vs. 221 (163 - 241), p=0.214] were shorter with HPC compared to non-HPC but without statistical significance. Total time post catheter use [HPC 90 s (69 - 105) vs. non-HPC 84 (70 - 97), p=0.496)] was similar for both. For participants with either tetra- and paraplegia as well as for HPC or non-HPC users, only step ‘prepare catheter’ was significantly shorter when using HPC compared to non-HPC (Table 4).

## DISCUSSION

To our knowledge, this is a novel cross-over RCT that quantifies the time taken to perform IC for individuals with SCI. A previous study examined preference and patient-reported time needed for catheterization across two coated catheters in a population of IC users^28^, but did not report quantitative measures of catheterization time, and examined two coated catheters in an overall IC population.

Twenty participants were timed while performing IC using HPCs and non-HPCs. Our results showed a substantial temporal burden from performing IC, independent of catheter type. The 90 second difference in median total time needed to perform IC between HPCs and non-HPC, corresponding to a 24% reduction in total time spent, was not only statistically significant but represents a meaningful difference. Considering an IC frequency of 4 to 6 times a day, those using HPCs will spend less than 24 minutes a day, while using non-HPCs will require around 31 minutes per day. This difference of 7 minutes per day adds up to 45 hours per year which roughly corresponds to one week of full-time employment. Arguably, this time difference may be important for all individuals, employers, and society. Even with HPCs being significantly faster to use, the temporal burden of IC remains significant regardless of catheter technology. As an illustrative example, an able-bodied person spends about 22 seconds^29^ to void. If we assume one person would spend the average time for each relevant step presented in table 2, this person would spend about 90 seconds per void or 7.5 minutes per day. In comparison, spending 24 to 31 minutes per day on IC clearly poses a significant temporal burden.

This substantial IC time use adds an additional item to the list of concerns faced by SCI individuals (including concerns such as difficulties finding accessible bathrooms, feeling of embarrassment and lack of privacy).^30^ In addition to posing an obstacle to maintaining efficient work hours^31^, the temporal burden may even exacerbate difficulties with adherence to IC as previously reported in the SCI population.^32^ Therefore, speed and efficiency of catheterization remains a high priority for individuals with SCI.

The difference in total time use across catheters was mainly attributed to the steps involving direct catheter use, while remaining steps had less impact overall (table 2). For example, there was a 63% reduction in median total time taken to prepare HPCs compared to non-HPCs while the time taken to exit the bathroom was similar between the catheters. This pattern applied collectively to all subgroups (i.e., individuals with tetraplegia or paraplegia, daily users of HPCs or non-HPCs) which may be explained by the properties of each catheter. For example, there is no requirement to manually lubricate HPCs. In addition, preparation of non-HPCs require individuals to open lubricant packaging and maintain at least a clean, if not sterile, technique until the insertion of the catheter. These requirements do not apply to HPCs.

Dexterity is an important factor that determines the difficulty of performing IC.^33^ Studies are still divided as to whether HPCs are easier or more difficult to handle.^34,35^ We hypothesized that individuals with tetraplegia would experience difficulties in handling HPCs due to impaired hand function compared to individuals with paraplegia. While both groups spent a similar time to prepare HPCs (16 vs 15 seconds), individuals with tetraplegia needed significantly more time (67 vs. 21 seconds, p=0.003) to prepare (i.e., unpack and lubricate) non-HPCs, than those with paraplegia.

Individuals rated HPCs to be easier to prepare than non-HPCs as well as causing less urethral burning sensation, which could be due to the need to lubricate non-HPCs prior to insertion. Even though there were no statistical differences (i.e., comparing the raw medians) in the remaining 11 satisfaction scores, HPCs scores were better in 6, while the remaining 5 were all tied. Several studies have reported participants finding HPCs easier to use compared to non-HPCs.^36,37^ To ensure that the results were not influenced by status quo bias, we split the sample analysis into subgroups depending on which type of catheter participants were regularly using (i.e., HPC vs non-HPC users). The results showed that no matter which type of catheter individuals were regularly using, participants catheterized faster with HPCs than with non-HPCs, driven by the preparation step.

While this study was intended to gain a first impression of the temporal burden associated with IC in individuals with SCI, the key limitation of this study was the relatively small sample size. Nevertheless, this study provides evidence that individuals with SCI performing IC could benefit from using HPCs with regard to a better user satisfaction and a lesser temporal burden. However, other factors playing significant roles in the lives of individuals with SCI must be considered. Ultimately, the ability for this cohort to use HPC depend on a host of factors, including coverage provided by the respective health care system, re-use (cost amplification associated with single use catheters)^38^ and environmental burden (i.e., ∼5 HPCs versus re-using 1 or 2 non-HPCs per day).^39^

## CONCLUSION

This pilot cross-over RCT provides evidence for a substantial IC time requirement for individuals with SCI, independent of the catheter type, with potential negative implications on levels of intra- and interpersonal stress, overall burden of SCI, workforce opportunities and IC compliance. The significance of reducing the time needed for IC while protecting the lower urinary tract makes it imperative that IC is performed efficiently which, in the context of this study, points to the merits of HPCs.

## Supporting information

Supplement Figure 1 - NON - Hydrophilic Catheter - Post-assessment Questionnaire

Supplement Figure 2 - Hydrophilic Catheter - Post-assessment Questionnaire

## Data Availability

Data are available from the authors upon request.

## ACKNOWLEDGMENTS

Andrei V. Krassioukov and Matthias Walter had full access to all the data in the study and take responsibility for the integrity of the data and the accuracy of the data analysis.

## Study concept and design

Karthik Gopalakrishnan, Nick Fabrin Nielsen, Andrea Ramirez, Jeppe Sørensen, Andrei V. Krassioukov, and Matthias Walter

## Generation of the random allocation sequence, enrollment of participants, and assignment of participants to interventions

Karthik Gopalakrishnan, Andrea Ramirez, and Matthias Walter

## Acquisition of data

Karthik Gopalakrishnan, Andrea Ramirez, Andrei V. Krassioukov, and Matthias Walter

## Drafting of the manuscript

Karthik Gopalakrishnan and Nick Fabrin Nielsen

## Critical revision of the manuscript for important intellectual content

Andrea Ramirez, Jeppe Sørensen, Andrei V. Krassioukov, and Matthias Walter

## Statistical analysis

Karthik Gopalakrishnan and Matthias Walter

## Supervision

Andrei V. Krassioukov and Matthias Walter

## Other

N/A.

