## Supplement Figure 1 - NON - Hydrophilic Catheter - Post-assessment Questionnaire for "Time needed to perform intermittent catheterization in adults with spinal cord injury: A pilot randomized controlled cross-over study"

**Assessment - PART 2**

**PARTICIPANT ID:** ____________________ **ASSESSOR INITIALS:** ____________________

**DATE:** **Month:____ Day:____ Year:_____**

### Questions related to your catheterization habits **Visit: ☐ 3 or ☐ 4** **Type (B)**

| | | |
|---|---|---|
| 1. Do you transfer to the toilet before your start catheterization? | ☐ Yes | ☐ No |
| 2. Do you clean your genitalia before your start catheterization? | ☐ Yes | ☐ No |
| 3. Do you have had a urinary tract infection over the last 12 months? | ☐ Yes | ☐ No |
| 4. Do you re-use catheter? | ☐ Yes | ☐ No |

### General questions

| This catheter is … | Strongly agree | Agree | Neutral | Disagree | Strongly disagree |
|---|---|---|---|---|---|
| easy to prepare | ☐ | ☐ | ☐ | ☐ | ☐ |
| easy to handle before insertion | ☐ | ☐ | ☐ | ☐ | ☐ |
| easy to insert | ☐ | ☐ | ☐ | ☐ | ☐ |
| able to drain my bladder completely | ☐ | ☐ | ☐ | ☐ | ☐ |
| easy to remove | ☐ | ☐ | ☐ | ☐ | ☐ |
| easy to dispose | ☐ | ☐ | ☐ | ☐ | ☐ |

### Catheter-specific questions

| | Strongly agree | Agree | Neutral | Disagree | Strongly disagree |
|---|---|---|---|---|---|
| I did not feel any pain during insertion | ☐ | ☐ | ☐ | ☐ | ☐ |
| I did not feel any burning sensation during insertion | ☐ | ☐ | ☐ | ☐ | ☐ |
| I don't like that I have to add lubrication | ☐ | ☐ | ☐ | ☐ | ☐ |
| The length of this catheter is sufficient to empty my bladder | ☐ | ☐ | ☐ | ☐ | ☐ |
| Overall, I am satisfied with this catheter | ☐ | ☐ | ☐ | ☐ | ☐ |
| I would recommend this catheter to someone else | ☐ | ☐ | ☐ | ☐ | ☐ |

### Only for the second assessment

| | Strongly agree | Agree | Neutral | Disagree | Strongly disagree |
|---|---|---|---|---|---|
| I would prefer this catheter over the one from the first assessment | ☐ | ☐ | ☐ | ☐ | ☐ |
